# Trends in cross-border and illicit tobacco purchases and associations with motivation to stop smoking and quit attempts: a representative survey of smokers in England, 2019-2022

**DOI:** 10.1101/2023.02.03.23285421

**Authors:** Sarah E. Jackson, Sharon Cox, Jamie Brown

**Affiliations:** Department of Behavioural Science and Health, University College London, London, UK; SPECTRUM Consortium, UK

**Keywords:** illicit tobacco, cross-border tobacco, tax avoidance, tax evasion, tobacco tax, motivation, quit attempt, smoking

## Abstract

**Objectives:** The last five years have seen substantial changes in England’s social and economic landscape as a result of Brexit, the Covid-19 pandemic, and cost-of-living crisis. We aimed to examine changes in cross-border and illicit tobacco purchasing, and associations with quitting activity, over this period.

**Design:** Nationally-representative monthly cross-sectional survey.

**Setting:** England, 2019-2022.

**Participants:** 11,232 adult (≥18y) past-year smokers.

**Main outcome measures:** We estimated time trends in the proportion of smokers reporting purchasing tobacco from (i) cross-border and (ii) illicit sources in the past 6 months, and examined associations with motivation to stop smoking (among current smokers) and past-year quit attempts (among past-year smokers).

**Results:** Between February 2019 and October 2022, there was a non-linear increase in the proportion of smokers reporting purchasing cross-border tobacco (from 5.2% to 16.1%; PR=3.10, 95%CI=2.03-4.73) but no overall change in the proportion reporting purchasing illicit tobacco (from 9.2% to 8.5%; PR=0.92, 95%CI=0.70-1.21). Both cross-border (OR_adj_=0.65, 95%CI=0.56-0.77) and illicit (OR_adj_=0.74, 95%CI=0.63-0.86) tobacco purchasing were associated with lower odds of reporting a recent quit attempt. Smokers who purchased cross-border tobacco also reported lower motivation to stop smoking (OR_adj_=0.84, 95%CI=0.75-0.95).

**Conclusions:** Despite a fall in cross-border tobacco purchasing during the first year of the Covid-19 pandemic, the proportion of smokers in England reporting purchasing cross-border tobacco is now three times higher than it was at the start of 2019. The proportion reporting purchasing illicit tobacco has not changed substantially. Tackling the increasing use of cheap tobacco in England may be an important target for motivating quit attempts.

**What this paper adds:** *What is already known on this topic:* Tobacco tax avoidance and evasion strategies, such as buying tobacco cheaply from cross-border or illicit sources, undermine the effectiveness of tax policy.

*What this study adds:* Since February 2019, the proportion of smokers in England reporting purchasing cross-border tobacco has tripled, while the proportion reporting purchasing illicit tobacco remains similar. Smokers who use cheap tobacco are less likely to try to quit.

*How this study might affect research, practice or policy:* Policy measures that reduce smokers’ access to these cheaper sources of tobacco could help to increase the rate of quit attempts among smokers and accelerate progress toward the government’s smokefree 2030 target.

## Introduction

Raising taxes on tobacco is an effective way to reduce smoking prevalence and tobacco consumption^1,2^ and reduce inequalities in smoking.^3,4^ Tobacco tax avoidance and tax evasion strategies undermine the effectiveness of tax policy by allowing smokers to buy tobacco at cheaper prices. Understanding how use of these strategies is changing over time, and what impact these changes have on quitting activity, is important for informing tobacco control policy.

Tax avoidance strategies include purchasing tobacco legally from low-tax jurisdictions across international borders, or duty-free while travelling between countries (‘cross-border purchases’).^5^ Tax evasion strategies include obtaining tobacco from illegal sources where no tax is paid at all, such as smuggling or buying counterfeit tobacco (‘illicit purchases’).^6,7^ Between 2002 and 2014, 12–20% of smokers in the UK reported having last purchased cigarettes from a low or untaxed source.^8^ The majority (≥75% in most years) of this group (8–16% of all smokers) reported cross-border purchases, with just 16–33% (2.6–3.7% of all smokers) buying from illicit sources (e.g., from informal sellers or friends/relatives).^8^

Several factors are likely to have affected the availability of, and smokers’ motivation to use, illicit and cross-border tobacco in England in recent years. First, as a result of Brexit, people travelling from the UK to countries within the European Union (EU) have been able to buy and bring back duty-free cigarettes since January 2021.^9^ Second, the Covid-19 pandemic (from March 2020) restricted social interaction and international travel, which may have reduced people’s access to cheap tobacco. Finally, the pandemic and, more recently, the ongoing cost-of-living crisis (since late 2020) have exposed many people to financial hardship as a result of loss of earnings^10^ and the cost of everyday essentials like groceries, household and energy bills rising faster than average household incomes.^11,12^ This may have increased smokers’ motivation to seek tobacco from low or untaxed sources to reduce the cost of smoking,^13,14^ particularly among less advantaged socioeconomic groups (e.g., those on a low income).^14,15^

By making smoking more affordable than other sources of purchase, any increase in cross-border or illicit tobacco purchasing may reduce smokers’ motivation to quit, undermining the effectiveness of tobacco tax policy for reducing smoking prevalence. Smokers who report buying cigarettes from low/untaxed sources – and those who switch to purchasing cheap tobacco – have been shown to be significantly less likely to make a quit attempt than those who continue to pay the full amount of tax.^16,17^ The extent to which the associations of cross-border and illicit tobacco purchasing with motivation to quit and quit attempts are moderated during a period of substantial change is not clear.

This study therefore aimed to examine changes in purchasing illicit and cross-border tobacco by smokers in England since 2019 (a year before the UK left the EU) and associations with changes in quitting activity. Specifically, we addressed the following research questions (RQs):

1. How have the proportions of past-year smokers in England self-reporting (i) purchasing cross-border tobacco and (ii) purchasing illicit tobacco changed between 2019 and 2022?
2. To what extent are cross-border and illicit tobacco purchasing associated with:
  a. Motivation to stop smoking among current smokers;
  b. Quit attempts among past-year smokers?
3. Has the magnitude of associations in RQ2 varied between 2019 and 2022?

For each RQ, we analysed data overall and by occupational social grade, to explore differences by socioeconomic position.

## Method

### Design

Data were drawn from the ongoing Smoking Toolkit Study, a monthly cross-sectional survey of a representative sample of adults in England designed to provide insights into population-wide influences on smoking.^18,19^ The study uses a hybrid of random probability and simple quota sampling to select a new sample of approximately 1,700 adults (≥18 years) each month. Full details of the sampling procedure are provided elsewhere.^19^

Data were collected monthly through face-to-face computer-assisted interviews up to February 2020. However, social distancing restrictions under the Covid-19 pandemic meant that no data were collected in March 2020, and data from April 2020 onwards have been collected via telephone. Comparisons of data collected face-to-face with sales data and other national surveys indicate that key variables such as key sociodemographic variables, smoking prevalence, and cigarette consumption are nationally representative.^18,20^ The telephone-based data collection relies upon the same combination of random location and quota sampling, and weighting approach as the face-to-face interviews and comparisons of the two data collection modalities indicate good comparability.^21–23^

For the present study, we used data collected from participants in the period from February 2019 (a year before the UK left the EU) to October 2022 (the most recent data available on source of purchase of tobacco at the time of analysis) who reported being a current smoker or having quit in the past year (‘past-year smokers’).

### Measures

***Smoking status*** was assessed with the question: ‘Which of the following best applies to you?

a. I smoke cigarettes (including hand-rolled) every day;
b. I smoke cigarettes (including hand-rolled), but not every day;
c. I do not smoke cigarettes at all, but I do smoke tobacco of some kind (e.g. pipe, cigar or shisha);
d. I have stopped smoking completely in the last year;
e. I stopped smoking completely more than a year ago;
f. I have never been a smoker (i.e. smoked for a year or more).’

Current smokers were those who responded *a* to *c*. Past-year smokers were those who responded *a* to *d*.

Where smokers purchase their tobacco was assessed with the question: ‘In the last 6 months, have you bought any cigarettes or hand rolled tobacco from any of the following?’ Participants could select multiple responses from a list of options. ***Cross-border purchasing*** was coded 1 for those who reported buying cigarettes or tobacco abroad and bringing them back with them, or friends/family bought from abroad, else it was coded 0. Duty free sources within the UK were not specified as a response option and some respondents may have included these in their definition of cross-border sources. ***Purchase from illicit sources*** was coded 1 for those who reported buying cigarettes or tobacco under the counter (from newsagent, off-license, or corner shop), in a pub (somebody comes around selling cheap), from people who sell cheap cigarettes on the street, from people in the local area who are a trusted source of cheap cigarettes, or cheap from friends, else it was coded 0. The item assessing source of purchase was included in all monthly waves up to April 2022, then reduced to quarterly assessment (July and October 2022) due to funding changes.

Among current smokers, ***motivation to stop smoking*** was assessed using the Motivation to Stop Scale.^24^ Smokers were asked: ‘Which of the following describes you?

1. I don’t want to stop smoking;
2. I think I should stop smoking but don’t really want to;
3. I want to stop smoking but haven’t thought about when;
4. I REALLY want to stop smoking but I don’t know when I will;
5. I want to stop smoking and hope to soon;
6. I REALLY want to stop smoking and intend to in the next 3 months;
7. I REALLY want to stop smoking and intend to in the next month.’

Motivation to stop smoking was analysed as an ordinal variable.

Among past-year smokers, ***quit attempts*** were assessed with the question: ‘How many serious attempts to stop smoking have you made in the last 12 months? By serious attempt I mean you decided that you would try to make sure you never smoked again.’ Those who reported having made at least one past-year quit attempt were coded 1 and those who reported no quit attempts were coded 0.

**Social grade** was categorised based on National Readership Survey classifications^25^ as ABC1, which includes managerial, professional, and upper supervisory occupations and C2DE, which includes manual routine, semi-routine, lower supervisory, and long-term unemployed.

Covariates for RQ2 and RQ3 included age, gender, and social grade.

### Statistical analysis

The analysis plan was pre-registered on Open Science Framework (https://osf.io/eapcw/). We made one amendment to the analysis plan: we had planned to analyse data up to December 2022 (the most recent data available at the time of analysis) but no data on source of purchase were collected in November or December, so were only able to include data up to October 2022. Data were analysed in R v.4.2.2. The Smoking Toolkit Study uses raking to match the sample to the population in England on the dimensions of age, social grade, region, housing tenure, ethnicity, and working status within sex.^26^ This profile is determined each month by combining data from the 2011 UK Census, the Office for National Statistics mid-year estimates, and the annual National Readership Survey.^18^ The following analyses were done using weighted data.

#### RQ1: Time trends

We used logistic regression to estimate monthly time trends in the proportion of past-year smokers purchasing (i) cross-border tobacco and (ii) illicit tobacco in the past 6 months. For the overall analysis, models only included time (survey month) as a predictor. For the social grade-specific analysis, models included time, social grade, and their interaction as predictors – this allows for time trends to differ across social grades. Survey month was modelled using restricted cubic splines, to allow relationships with time to be flexible and non-linear, while avoiding categorisation. We had planned to use three knots placed at the earliest, middle, and latest months, but visual inspection of the modelled estimates against raw quarterly data points indicated the model did not provide a good fit for trends in cross-border tobacco purchasing. We therefore reran the models for both cross-border and illicit tobacco purchasing using four knots and compared model fit using the Akaike information criterion (AIC; see Supplementary File). The criteria for selecting the best fitting model was either the model with the lowest AIC or the simplest model if it was within two units of the model with the lowest AIC score. Our interpretation was based on the best fitting model for each outcome: four knots for cross-border tobacco purchasing and three knots for illicit tobacco purchasing. Prevalence ratios for changes in prevalence across the whole time-series (October 2022 versus February 2019) are presented, alongside 95% CIs calculated using bootstrapping.

#### RQ2 and RQ3: Associations with quitting activity

We assessed associations of (i) purchasing cross-border tobacco and (ii) purchasing illicit tobacco with (a) motivation to quit using ordinal logistic regression and (b) quit attempts using binary logistic regression. For each association, we ran a series of four models: Model 1 was unadjusted; Model 2 was adjusted for covariates (age, gender, and social grade); Model 3 tested for moderation by social grade, adding the two-way interaction between purchasing cross-border/illicit tobacco and social grade to Model 2; and Model 4 tested for moderation by survey year (RQ3), adding the two-way interaction between purchasing cross-border/illicit tobacco and survey year (2019-2022, analysed as a categorical variable) to Model 2.

## Results

A total of 71,993 adults aged ≥18 were surveyed in England between February 2019 and October 2022, of whom 12,432 (17.3%) were past-year smokers. We excluded 1,200 surveyed in waves that did not assess source of purchase of tobacco (May/June/August/September 2022), leaving a final sample for analysis of 11,232 past-year smokers (46.2% female; mean [SD] age 41.8 [16.7] years; 58.4% social grade C2DE), of whom 10,073 were current smokers (46.0% female; mean [SD] age 42.1 [16.7] years; 59.5% social grade C2DE).

**Table 1** provides descriptive data on cross-border and illicit tobacco purchasing and **Figure 1** shows modelled estimates. Across the study period, 9.6% [95%CI 9.01-10.2%] of past-year smokers in England reported having purchased cross-border tobacco in the past 6 months and 10.3% [9.7-10.9%] reported having purchased illicit tobacco. Cross-border tobacco purchasing was less common among smokers from social grades C2DE compared with ABC1 (7.6% [6.9-8.3%] vs. 11.6% [10.8-12.4%]) but illicit tobacco purchasing was more common (11.8% [11.0-12.7%] vs. 8.8% [8.1-9.6%]).

**Table 1.** Trends in cross-border and illicit tobacco purchasing prevalence among past-year smokers in England

|  | Prevalence |  |  | Prevalence ratio<br>Feb 19 – Oct 22<br>[95% CI] |
| --- | --- | --- | --- | --- |
|  | Overall<sup>1</sup> | February<br>2019<sup>2</sup> | October<br>2022<sup>2</sup> |  |
| Purchased cross-border tobacco in past 6 months |  |  |  |  |
| All past-year smokers | 9.6% | 5.2% | 16.1% | 3.10 [2.03-4.73] |
| ABC1 | 11.6% | 6.6% | 23.3% | 3.52 [2.05-5.91] |
| C2DE | 7.6% | 4.4% | 11.5% | 2.61 [1.17-5.20] |
| Purchased illicit tobacco in past 6 months |  |  |  |  |
| All past-year smokers | 10.3% | 9.2% | 8.5% | 0.92 [0.70-1.21] |
| ABC1 | 8.8% | 6.8% | 6.8% | 1.01 [0.69-1.44] |
| C2DE | 11.8% | 10.6% | 9.9% | 0.93 [0.65-1.31] |

<sup>1</sup> Weighted prevalence based on data aggregated across the study period.
<sup>2</sup> Weighted prevalence from logistic regression on all past-year smokers and allowing an interaction between social grade and month (estimates for smokers from social grades ABC1 and C2DE), modelled non-linearly using restricted cubic splines (four knots for cross-border tobacco and three knots for illicit tobacco; see Supplementary File for details of model selection).

**Figure 1.**
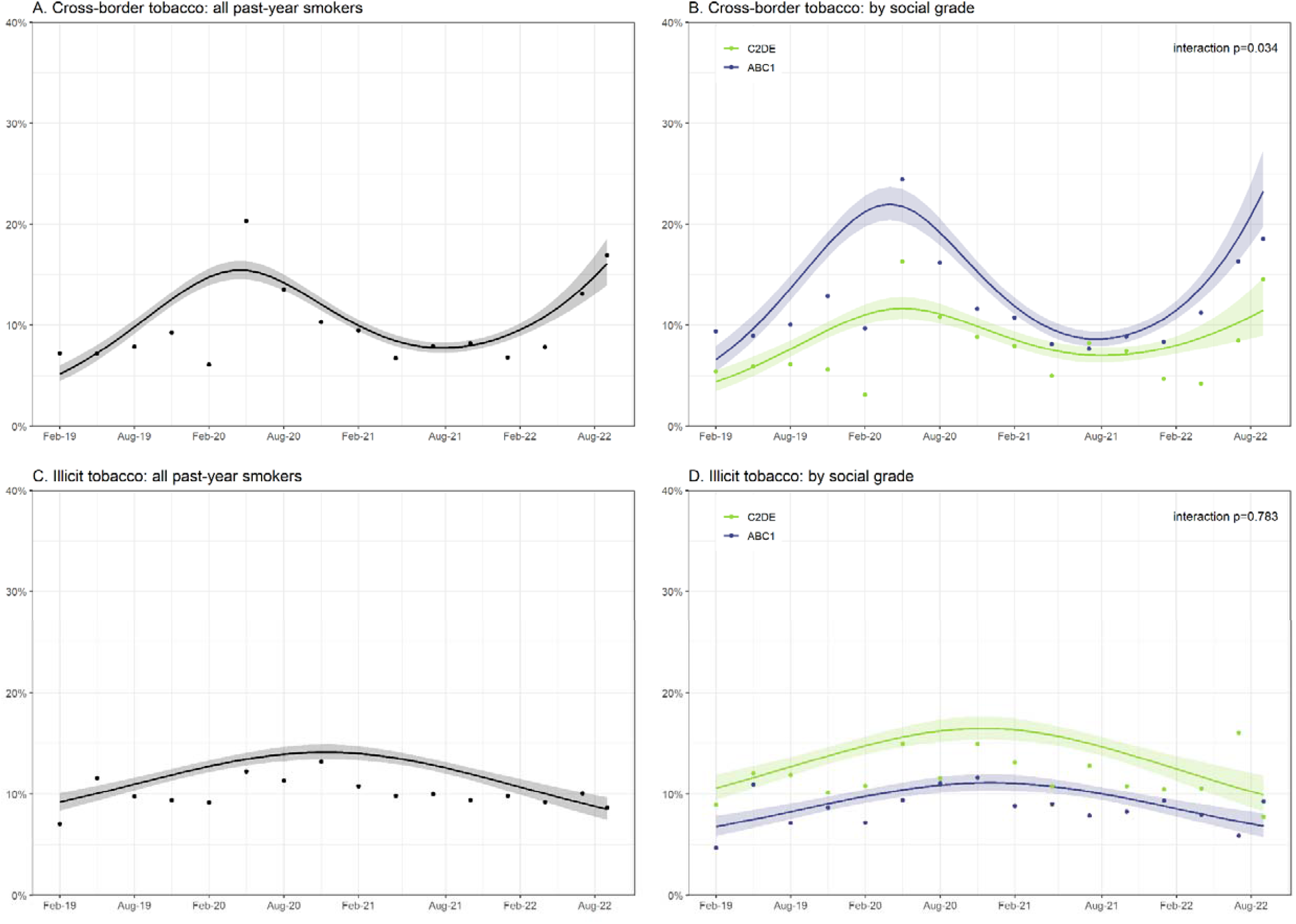
Percentage of past-year smokers purchasing cross-border and illicit tobacco in England, February 2019 to October 2022. Data are presented for all past-year smokers (left panel) and by social grade (right panel). Lines represent point estimates from logistic regression with survey month modelled non-linearly using restricted cubic splines (with four knots for cross-border tobacco and three knots for illicit tobacco; see Supplementary File for details of model selection). Shaded areas represent standard errors. Points represent raw weighted prevalence by quarter.

From February 2019 to October 2022, the proportion of smokers reporting having purchased cross-border tobacco increased from 5.2% to 16.1% (**Table 1**; PR=3.10, 95%CI 2.03-4.73). The increase over time was not linear: past-6-month cross-border tobacco purchasing increased from 5.2% to 15.4% between February 2019 and April 2020, fell to 7.8% between April 2020 and September 2021, and then increased to 16.1% by October 2022 (**Figure 1A**). Although an overall increase in prevalence of cross-border tobacco purchasing was observed across social grades (**Table 1**), time trends differed significantly within this period (interaction *p*=0.034), with more pronounced changes observed among smokers from social grades ABC1 than C2DE (**Figure 1B**).

The proportion of smokers reporting having purchased illicit tobacco did not change significantly from February 2019 to October 2022 (**Table 1;** PR=0.92, 95%CI 0.70-1.21), with prevalence rising from 9.2% to 14.2% between February 2019 and November 2020, then falling to 8.5% by October 2022 (**Figure 1C**). Trends in illicit tobacco purchasing did not differ significantly by social grade (**Table 1**; **Figure 1D**).

**Tables 2 and 3** summarise associations of quitting activity with cross-border and illicit tobacco purchasing, respectively. After adjustment for covariates (Model 2), smokers who reported purchasing cross-border tobacco had lower motivation to stop smoking (OR=0.84, 95%CI 0.75-0.95) and lower odds of having tried to quit in the past year (25.7% vs. 35.3%; OR=0.65, 95%CI 0.56-0.77; **Table 2**). Smokers who reported purchasing illicit tobacco had similar levels of motivation to stop smoking to those who did not (OR=0.93, 95%CI 0.83-1.05) but had lower odds of having tried to quit in the past year (30.7% vs. 34.8%; OR=0.74, 95%CI 0.63-0.86; **Table 3**). Associations did not differ significantly by social grade (Model 3) or survey year (Model 4), with the exception of the association between illicit tobacco purchasing and quit attempts, which differed significantly by survey year. In 2019-2021, smokers who reported purchasing illicit tobacco had lower odds of having made a quit attempt, whereas in 2022 they had higher odds (**Figure 2**).

**Table 2.**
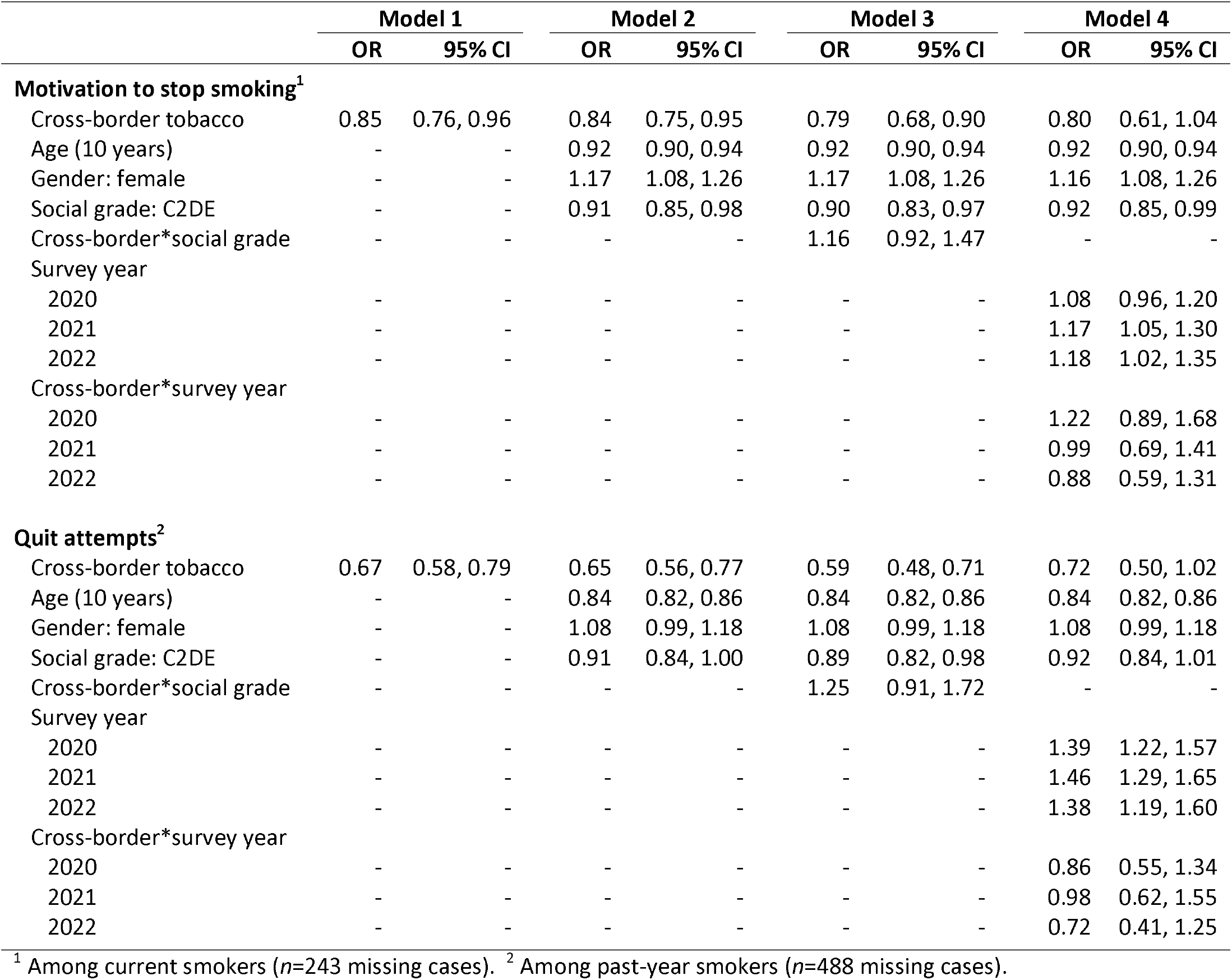
Associations between cross-border tobacco purchasing and quitting activity among smokers in England

|  | Model 1 |  | Model 2 |  | Model 3 |  | Model 4 |  |
| --- | --- | --- | --- | --- | --- | --- | --- | --- |
|  | OR | 95% CI | OR | 95% CI | OR | 95% CI | OR | 95% CI |
| **Motivation to stop smoking<sup>1</sup>** |  |  |  |  |  |  |  |  |
| Cross-border tobacco | 0.85 | 0.76, 0.96 | 0.84 | 0.75, 0.95 | 0.79 | 0.68, 0.90 | 0.80 | 0.61, 1.04 |
| Age (10 years) | - | - | 0.92 | 0.90, 0.94 | 0.92 | 0.90, 0.94 | 0.92 | 0.90, 0.94 |
| Gender: female | - | - | 1.17 | 1.08, 1.26 | 1.17 | 1.08, 1.26 | 1.16 | 1.08, 1.26 |
| Social grade: C2DE | - | - | 0.91 | 0.85, 0.98 | 0.90 | 0.83, 0.97 | 0.92 | 0.85, 0.99 |
| Cross-border*social grade | - | - | - | - | 1.16 | 0.92, 1.47 | - | - |
| Survey year |  |  |  |  |  |  |  |  |
| 2020 | - | - | - | - | - | - | 1.08 | 0.96, 1.20 |
| 2021 | - | - | - | - | - | - | 1.17 | 1.05, 1.30 |
| 2022 | - | - | - | - | - | - | 1.18 | 1.02, 1.35 |
| Cross-border*survey year |  |  |  |  |  |  |  |  |
| 2020 | - | - | - | - | - | - | 1.22 | 0.89, 1.68 |
| 2021 | - | - | - | - | - | - | 0.99 | 0.69, 1.41 |
| 2022 | - | - | - | - | - | - | 0.88 | 0.59, 1.31 |
| **Quit attempts<sup>2</sup>** |  |  |  |  |  |  |  |  |
| Cross-border tobacco | 0.67 | 0.58, 0.79 | 0.65 | 0.56, 0.77 | 0.59 | 0.48, 0.71 | 0.72 | 0.50, 1.02 |
| Age (10 years) | - | - | 0.84 | 0.82, 0.86 | 0.84 | 0.82, 0.86 | 0.84 | 0.82, 0.86 |
| Gender: female | - | - | 1.08 | 0.99, 1.18 | 1.08 | 0.99, 1.18 | 1.08 | 0.99, 1.18 |
| Social grade: C2DE | - | - | 0.91 | 0.84, 1.00 | 0.89 | 0.82, 0.98 | 0.92 | 0.84, 1.01 |
| Cross-border*social grade | - | - | - | - | 1.25 | 0.91, 1.72 | - | - |
| Survey year |  |  |  |  |  |  |  |  |
| 2020 | - | - | - | - | - | - | 1.39 | 1.22, 1.57 |
| 2021 | - | - | - | - | - | - | 1.46 | 1.29, 1.65 |
| 2022 | - | - | - | - | - | - | 1.38 | 1.19, 1.60 |
| Cross-border*survey year |  |  |  |  |  |  |  |  |
| 2020 | - | - | - | - | - | - | 0.86 | 0.55, 1.34 |
| 2021 | - | - | - | - | - | - | 0.98 | 0.62, 1.55 |
| 2022 | - | - | - | - | - | - | 0.72 | 0.41, 1.25 |

<sup>1</sup> Among current smokers ( $n=243$ missing cases). <sup>2</sup> Among past-year smokers ( $n=488$ missing cases).

**Table 3.**
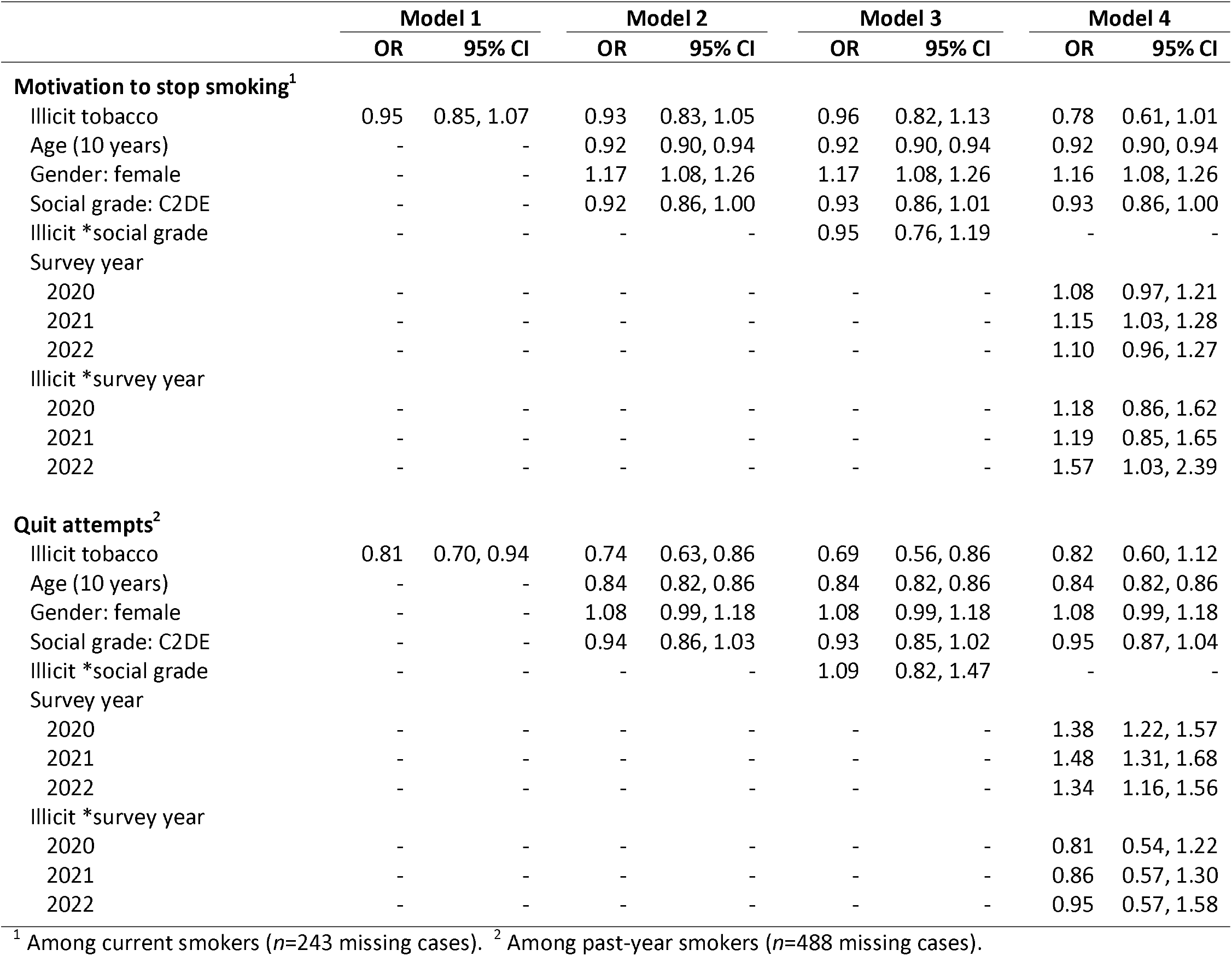
Associations between illicit tobacco purchasing and quitting activity among smokers in England

**Figure 2.**
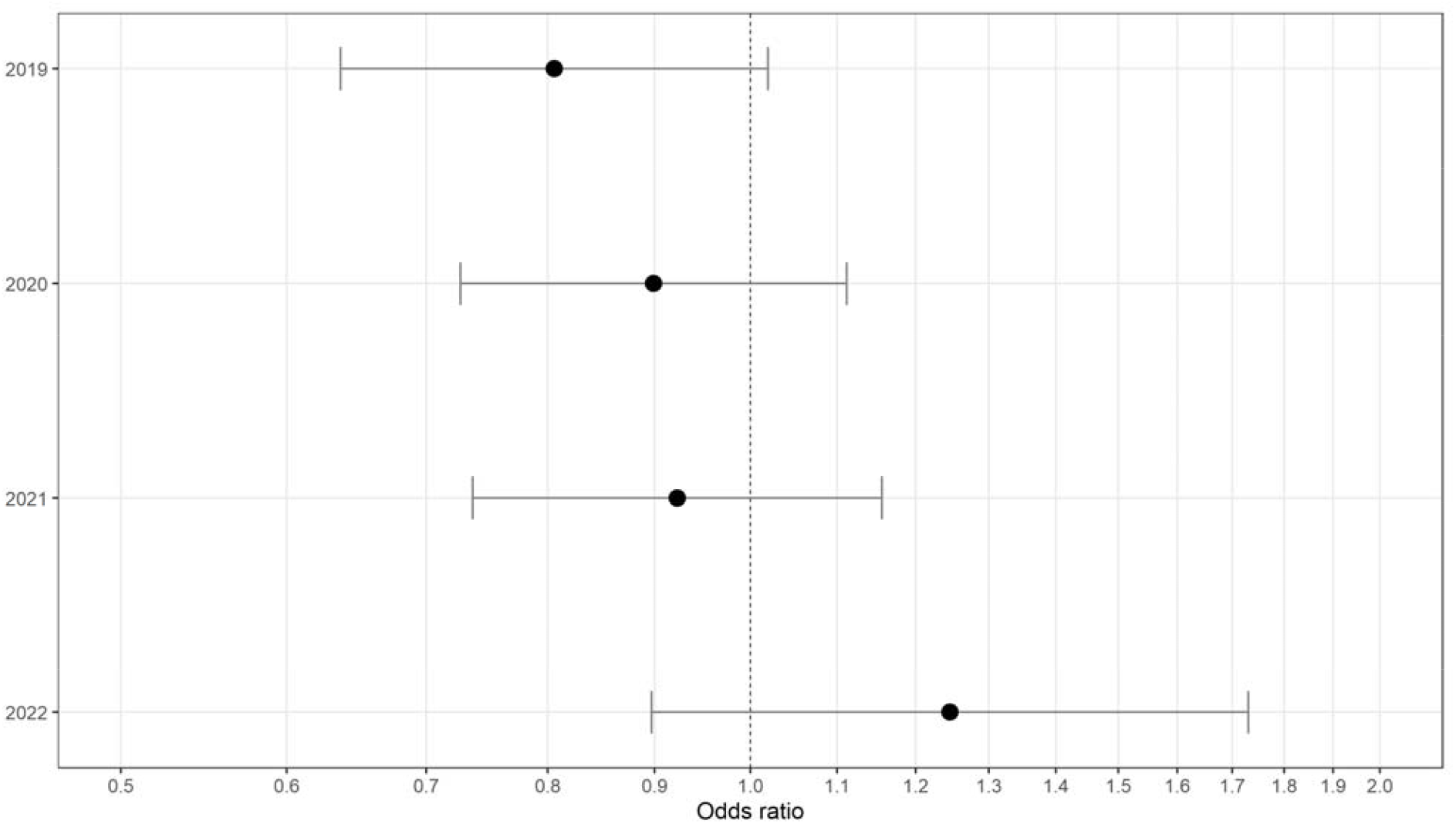
Association between illicit tobacco purchasing and motivation to stop smoking by survey year. Points represent odds ratios from adjusted logistic regression models stratified by survey year. Error bars represent 95% confidence intervals.

## Discussion

Between February 2019 and October 2022, there was a non-linear increase in the proportion of smokers in England reporting purchasing cross-border tobacco but no overall change in the proportion reporting purchasing illicit tobacco. Both cross-border and illicit tobacco purchasing were associated with lower odds of reporting a recent quit attempt. Smokers who purchased cross-border tobacco also reported lower motivation to stop smoking.

The curvilinear trend in cross-border tobacco purchasing might be explained by changes in motivation and access resulting from Brexit and the Covid-19 pandemic. Reports of purchasing cross-border tobacco in the past 6 months tripled between February 2019 and April 2020, with the raw data points indicating a sharp rise in April 2020. It is possible this was due to people thinking cross-border tobacco would be cheaper as a result of the UK leaving the EU in January 2020 (despite EU duty-free purchasing not being implemented until the end of the transition period in January 2021^9^). Alternatively, it could be that smokers who were travelling in January-March 2020 thought it might be wise to stock up as it became evident that Covid-19 was coming to the UK and was likely to affect future travel plans. After the Covid-19 pandemic reached the UK and restrictions on international travel were implemented, past-6-month cross-border tobacco purchases declined substantially, then rebounded rapidly from September 2021 as people began travelling abroad again over the summer.^27^ The prevalence of cross-border tobacco purchasing was higher, and changes over time were more pronounced, among smokers from more versus less advantaged social grades. This is consistent with advantaged groups being more likely to travel by air than those with lower incomes,^28^ providing greater opportunity to purchase cheaper tobacco abroad.

Illicit tobacco purchasing showed less variability over time, rising by around 50% between February 2019 and November 2020 and returning to baseline levels by October 2022. This suggests that (i) the Covid-19 pandemic and associated restrictions on social interaction did not substantially reduce smokers’ access to illicit tobacco and (ii) the proportion of smokers buying tobacco from illicit sources has not (yet) increased during the cost-of-living crisis since late 2021 through to October 2022. While prevalence of illicit tobacco purchasing was higher among smokers from less versus more advantaged social grades, time trends were similar, showing no evidence of increased use of illicit tobacco among those with lower disposable incomes as economic pressures have heightened.

Consistent with previous findings,^17^ we observed lower odds of quit attempts among smokers who reported purchasing cross-border or illicit tobacco. This occurred across social grades and survey years, with the exception of 2022, when the point estimate of the association between illicit tobacco purchasing and quit attempts was reversed, although the confidence interval included no association and values consistent with the other years. It is not clear why this may have occurred and continued monitoring may provide further insight. Nevertheless, our findings collapsed across the period suggest smokers who use cheap tobacco are less likely to try to quit. This underscores the potential for cross-border and illicit tobacco to undermine the effectiveness of tax policy for reducing smoking prevalence. Policy measures that reduce smokers’ access to these cheaper sources of tobacco could help to increase the rate of quit attempts among smokers and accelerate progress toward the government’s smokefree 2030 target. Recent reductions in the budgets of trading standards in England have limited their capacity to tackle illicit tobacco.^29^ These should be reversed and dedicated to enforcement activity in local areas. A low-cost tobacco retailer registration scheme with sanctions could be implemented.^29,30^ The sanctions could provide additional funds for enforcement, and detailed surveillance of legal tobacco retailers.

Strengths of this study include the large, representative sample and the repeat cross-sectional design pre-dating Brexit and the Covid-19 pandemic. There were also limitations. Data on cross-border and illicit tobacco purchasing were self-reported. It is possible that participants may under-report the purchase of illicit tobacco due to legal concerns, which may underestimate its prevalence. Measures asked about tobacco purchasing over the past 6 months, introducing scope for recall bias. While this might have resulted in prevalence being underestimated, it should not have affected time trends as there is no reason to believe that recall would have changed over time. Another limitation was the change in modality of data collection from face-to-face (up to February 2020) to telephone interviews (from April 2020). While this was unavoidable due to social distancing restrictions implemented during the Covid-19 pandemic, it is possible that it contributed to some of the changes we observed; for example, the spike in reports of cross-border tobacco purchasing in the spring of 2020. However, the fact that we did not see a comparable change in reports of illicit tobacco purchasing does not point to there having been an impact on responses to the source of purchase question (such as participants being more likely to report purchasing cheap tobacco via telephone compared with face-to-face). In addition, comparisons of the face-to-face and telephone data within the Smoking Toolkit Study,^23^ combined with previous studies showing a high degree of comparability between face-to-face and telephone interviews,^31,32^ suggest it is reasonable to compare data collected via the two methods. Finally, while the sample was nationally representative, participants were recruited from households, meaning people experiencing homelessness – who have much higher smoking prevalence^33^ and who regularly smoke illicit tobacco^34^ – are not captured, which may underestimate illicit tobacco purchasing.

In conclusion, despite a fall in cross-border tobacco purchasing during the first year of the Covid-19 pandemic, the proportion of smokers in England reporting purchasing cross-border tobacco is now three times higher than it was at the start of 2019. The proportion reporting purchasing illicit tobacco has not changed substantially. Given smokers who reported purchasing cross-border and illicit tobacco reported lower levels of quitting activity, tackling increasing use of cheap tobacco may be an important target for motivating quit attempts.

## Data Availability

All data produced in the present study are available upon reasonable request to the authors

## Declarations

### Ethics approval

Ethical approval for the STS was granted originally by the UCL Ethics Committee (ID 0498/001). The data are not collected by UCL and are anonymized when received by UCL.

