## Supplementary material for "Trends in cross-border and illicit tobacco purchases and associations with motivation to stop smoking and quit attempts: a representative survey of smokers in England, 2019-2022"

**Supplementary File**

**Figure S1.** Percentage of past-year smokers purchasing cross-border and illicit tobacco in England, February 2019 to October 2022: modelled using restricted cubic splines with three knots

**Figure S2.** Percentage of past-year smokers purchasing cross-border and illicit tobacco in England, February 2019 to October 2022: modelled using restricted cubic splines with four knots

**Table S1.**  Comparison of model fit: three versus four knots

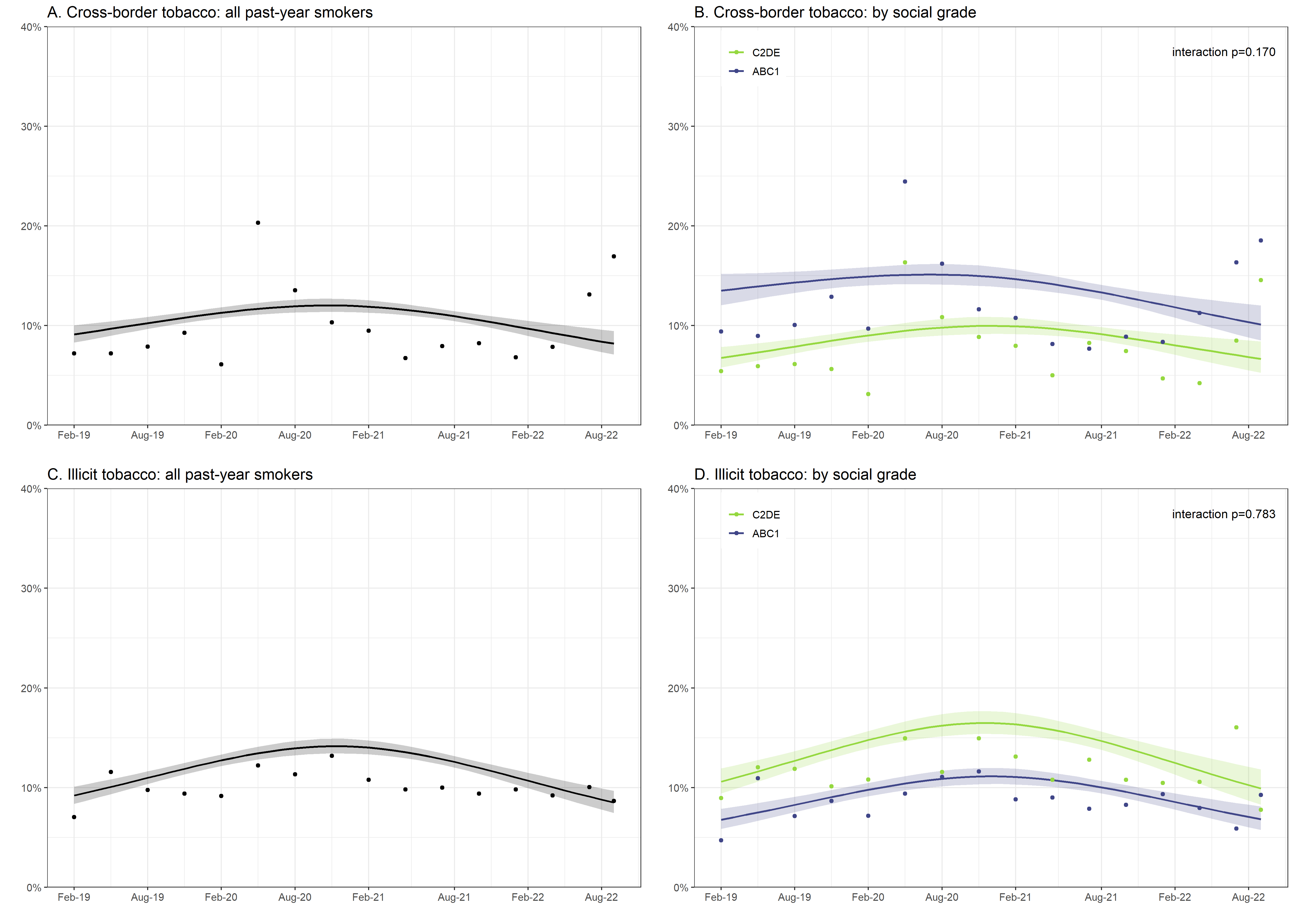

**Figure S1. Percentage of past-year smokers purchasing cross-border and illicit tobacco in England, February 2019 to October 2022: modelled using restricted cubic splines with three knots.** Data are presented for all past-year smokers (left panel) and by social grade (right panel). Lines represent point estimates from logistic regression with survey month modelled non-linearly using restricted cubic splines (three knots). Shaded areas represent standard errors. Points represent raw weighted prevalence by quarter.

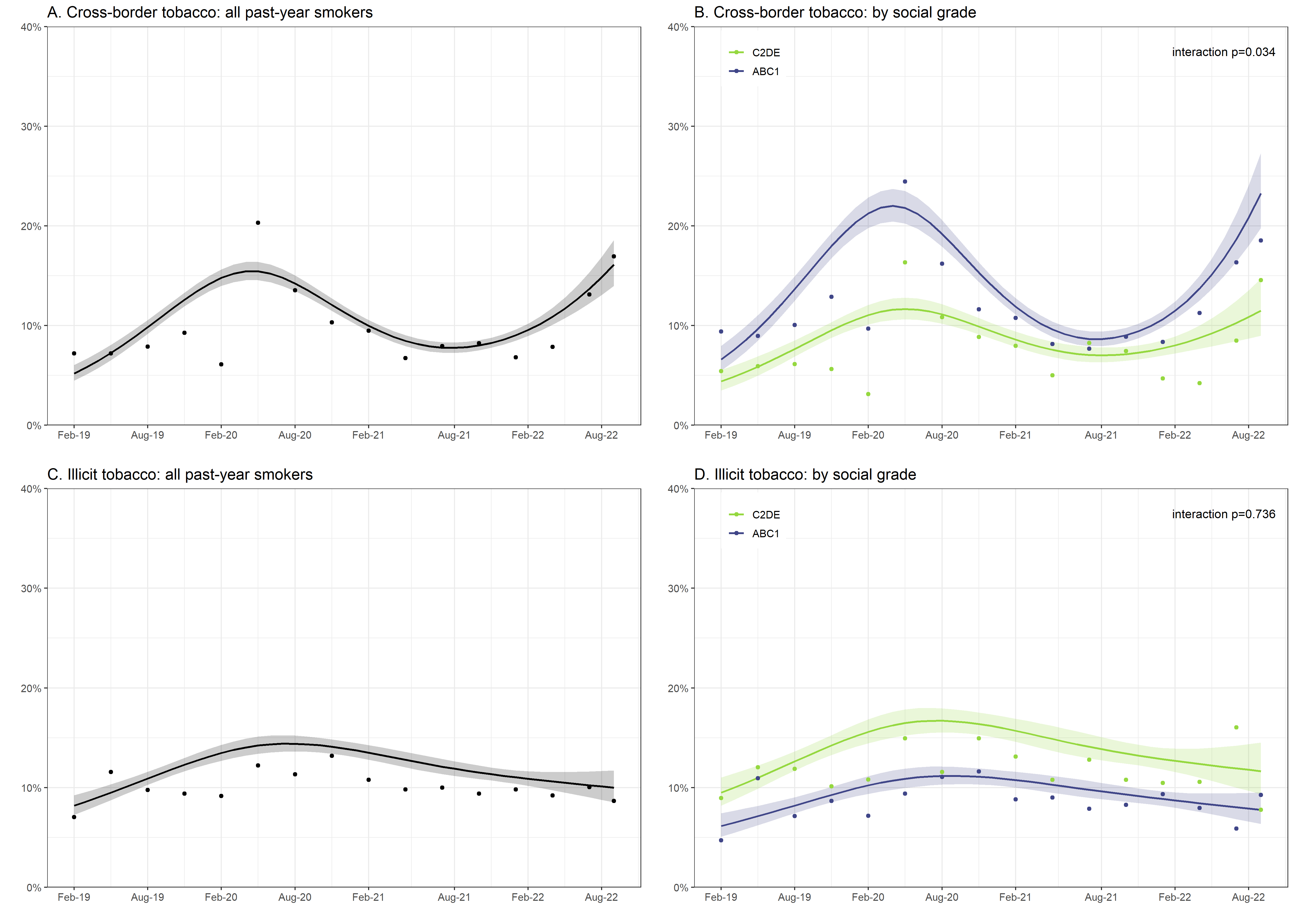

**Figure S2. Percentage of past-year smokers purchasing cross-border and illicit tobacco in England, February 2019 to October 2022: modelled using restricted cubic splines with four knots.** Data are presented for all past-year smokers (left panel) and by social grade (right panel). Lines represent point estimates from logistic regression with survey month modelled non-linearly using restricted cubic splines (three knots). Shaded areas represent standard errors. Points represent raw weighted prevalence by quarter.

| **Table S1.** Comparison of model fit: three versus four knots | | |  |
| --- | --- | --- | --- |
|  | **AIC** | | |
|  | **3 knots** | **4 knots** | **Difference** |
| Cross-border tobacco, all past-year smokers | 7167.83 | 7100.86 | -66.97 |
| Cross-border tobacco, by social grade | 7122.15 | 7050.47 | -71.68 |
| Illicit tobacco, all past-year smokers | 7686.10 | 7685.59 | -0.51 |
| Illicit tobacco, by social grade | 7652.85 | 7654.91 | 2.06 |
| AIC, Akaike information criterion. Lower values of AIC indicate better model fit. The criteria for selecting the best fitting model was either the model with the lowest AIC or the simplest model if it was within two units of the model with the lowest AIC score. | | | |
